# Performance of Nursing Activities Score in intensive care units of a teaching hospital in low middle income country: An observational study

**DOI:** 10.64898/2026.03.10.26348109

**Authors:** Raj Kumar Mehta, Hafizah Che Hassan, Bishnu Bista, Mamata Sharma Neupane

## Abstract

**Background:** Nursing workload in intensive care units (ICUs) plays a crucial role in determining patient outcomes, quality of care, and healthcare system efficiency. The Nursing Activities Score (NAS) is a validated tool used internationally to measure nursing workload and estimate the proportion of nursing time required for patient care. However, evidence regarding its application in low- and middle-income countries (LMICs) remains limited. High nursing workload has been associated with increased mortality, prolonged ICU stay, and compromised patient safety. This study aimed to assess nursing workload using NAS in an ICU of a teaching hospital and evaluate its predictive ability for patient outcomes.

**Methods:** This observational study included 501 ICU patients admitted to a teaching hospital. NAS scores were recorded for each patient, and outcomes were categorized as survivors and non-survivors. The predictive ability of NAS was evaluated using Receiver Operating Characteristic (ROC) curve analysis. Model calibration was assessed using the Hosmer–Lemeshow goodness-of-fit test. Logistic regression analysis was performed to determine the association between NAS and mortality risk. The Mann–Whitney U test was used to compare NAS scores between survivors and non-survivors.

**Results:** The median NAS score was 83.40 (IQR: 101.10–68.30; range: 39.2–134.4), indicating a high level of nursing workload in the ICU. ROC analysis showed that NAS had good predictive ability for patient outcomes with an AUROC of 0.838 (p < 0.001). The optimal cut-off value was 90.40, with 73.5% sensitivity and 73.1% specificity. The Hosmer–Lemeshow test (p = 0.422) indicated good model fit. Logistic regression analysis showed that higher NAS scores were significantly associated with increased mortality risk (Exp(B) = 0.937, p < 0.001). Non-survivors had significantly higher NAS scores (110.70) compared with survivors (76.20, p < 0.001).

**Conclusion:** NAS is a reliable tool for assessing ICU nursing workload and predicting patient outcomes. Higher NAS scores reflect greater patient severity and increased risk of mortality, highlighting the importance of optimized staffing and workload management in ICU settings.

**Author Summary:** Intensive care units (ICUs) care for the most critically ill patients and require constant monitoring and complex nursing interventions. However, in many low- and middle-income countries, including Nepal, the number of available nurses is often insufficient compared with the high demand for intensive care services. This imbalance can increase nursing workload and may affect the quality and safety of patient care. Therefore, reliable tools are needed to measure nursing workload and help hospitals plan staffing more effectively.

This study evaluated the **Nursing Activities Score (NAS)**, a standardized tool used internationally to measure nursing workload, in the ICU of a teaching hospital. Data from **501 ICU patients** were analyzed to determine the level of nursing workload and whether NAS could predict patient outcomes. The findings showed that the nursing workload was high, with a median NAS score of **83.4**, indicating substantial nursing care requirements. Patients who did not survive had significantly higher NAS scores compared with survivors. NAS also showed good accuracy in predicting patient outcomes.

These findings suggest that NAS is a useful tool for measuring nursing workload and identifying critically ill patients who require more intensive care. Using NAS in ICUs may help hospitals optimize staffing, improve patient safety, and support better critical care management in resource-limited settings.

## 1. INTRODUCTION

Nursing workload in intensive care units (ICUs) is a crucial factor influencing patient safety, quality of care, and nurse well-being. The complexity of care in ICUs demands continuous monitoring, invasive procedures, medication administration, and emergency interventions, making nurse staffing and workload key determinants of patient outcomes [1,2]. Nurse staffing shortages and excessive workload have been associated with higher mortality rates, increased nosocomial infections, longer ICU stays, and increased burnout among nurses [3,4]. A study by Kartik et al. (2017) reported that nursing staff allocation is a major problem in many ICUs and hospitals in India [5]. Patil et al. (2023) further emphasized that a higher mortality rate found in an ICU of a public teaching hospital in India and attributed this finding to the low nurse-to-patient ratio due to nurse shortage, budgetary constraints, and higher nursing workload [6]. However, in low-middle-income countries (LMICs) like Nepal, meeting the recommended 1:1 nurse-to-patient ratio remains a significant challenge [7]. Consequently, nurses often manage multiple critically ill patients simultaneously, leading to increased workload, compromised patient safety, and suboptimal adherence to clinical protocols [8]. Adequate nurse staffing is essential for maintaining high-quality patient care in ICUs. Studies indicate that higher nurse-patient ratios lead to better patient outcomes, while staff shortages increase the risk of mortality and medical errors [1,9]. A systematic review by Kane et al. (2007) confirmed that an increase in registered nurse staffing is associated with lower hospital mortality, reduced infections, and improved recovery rates [4]. Similarly, Numata et al. (2006) found that ICUs with lower nurse-to-patient ratios had higher mortality rates, underscoring the importance of appropriate staffing in critical care settings [10]. In many LMICs, the ICU nurse-to-patient ratio often exceeds the recommended standards, sometimes reaching 1:3 or more due to staff shortages and limited resources [7]. This excessive workload not only increases the risk of burnout among nurses but also jeopardizes patient safety and care quality [8]. Park J et al. (2022) reported that excessive nursing workload leads to cognitive overload, which can impair clinical decision-making and increase the likelihood of medical errors [2]. In such settings, tools for objectively measuring nursing workload are essential for workforce management and quality improvement initiatives. The Nursing Activities Score (NAS) is a validated tool used to measure nursing workload in ICUs. Developed by Miranda et al. (2003), the NAS is widely used in high-income countries to quantify the time and effort required for nursing care in critically ill patients [11]. The NAS assigns numerical values to different nursing activities, providing an objective measure of workload based on patient needs. Studies have shown that higher NAS scores correlate with increased nursing workload and patient severity [12]. However, despite its reliability, NAS has not been widely studied in LMICs, including Nepal, where ICU conditions and staffing challenges differ significantly from those in high-income countries. A study by Oliveira AC et al. (2016) found that NAS is a strong predictor of nursing workload, with higher NAS scores associated with increased nursing interventions and patient complexity [12]. Additionally, Carayon P et al. (2014) emphasized that measuring nursing workload is essential for maintaining patient safety, as excessive workload can lead to nurse fatigue, reduced vigilance, and an increased risk of errors. While NAS has been validated in various ICU settings, its application in resource-limited environments remains unexplored, highlighting a crucial knowledge gap in workload assessment in Nepalese ICUs [7]. A growing body of research links higher nursing workload to increased patient mortality. West et al. (2014) demonstrated that higher ICU nurse staffing levels were associated with lower mortality rates, suggesting that adequate nurse staffing is a protective factor against adverse outcomes [9]. Numata et al. (2006) further confirmed that ICU mortality rates are significantly influenced by nurse workload, particularly in high-acuity patients requiring intensive interventions [10]. Additionally, nursing workload has been associated with an increased risk of adverse events, including nosocomial infections, pressure ulcers, medication errors, and delayed interventions [4]. In ICUs with high nurse workload, infection rates are significantly higher, as nurses have limited time for infection control practices, such as hand hygiene and sterile procedures [12]. Furthermore, burnout and fatigue resulting from excessive workload can lead to decreased attentiveness and a higher likelihood of clinical errors. These findings underscore the urgent need for systematic workload assessments to optimize staffing and improve patient safety [2]. Despite global evidence on the importance of measuring nursing workload in ICUs, no study has been conducted in Nepal to assess nursing workload using the NAS tool. Given the staffing shortages, high patient burden, and limited resources in Nepalese hospitals, understanding ICU workload is critical for improving care quality and patient outcomes. The lack of objective workload assessments makes it difficult to identify staffing needs, optimize workforce distribution, and develop strategies to enhance patient safety [8]. The increasing demand for intensive care services in LMICs like Nepal, combined with nurse staffing shortages and excessive workload, highlights the need for systematic workload measurement tools. The NAS is a reliable and validated tool that can provide critical insights into ICU nursing workload, allowing for better resource allocation and workforce planning. Given the lack of research on NAS application in Nepal, this study is expected to fill an important knowledge gap by providing data-driven insights into ICU nursing workload and its impact on patient outcomes. By implementing evidence-based staffing models, hospitals can improve ICU efficiency, nurse well-being, and overall patient safety. This study aims to assess the nursing workload in Nepalese ICUs using the NAS and evaluate its predictive ability for patient outcomes. By analyzing the relationship between NAS scores and ICU mortality rates, this research seeks to provide evidence-based recommendations for staffing optimization and improving patient care in resource-limited settings.

## 2. MATERIALS AND METHODS

This study utilized a prospective observational design to assess nursing workload using the Nursing Activities Score (NAS) and evaluate its predictive ability for patient outcomes in an intensive care unit (ICU) of a teaching hospital in a low-middle-income country (LMIC). The study was conducted in the ICU of a tertiary care teaching hospital where critically ill patients receive continuous nursing and medical care. The ICU follows a standardized care protocol, with varying nurse-to-patient ratios depending on patient acuity. Inclusion Criteria were patients aged ≥18 years admitted to the ICU, patients requiring nursing interventions and continuous monitoring and patients who had at least 24 hours of ICU stay to ensure sufficient data collection. Exclusion Criteria were patients discharged or deceased within 24 hours of admission, patients transferred from or to other hospital units before 24 hours and cases with insufficient or missing data on NAS. A total of 501 ICU patients were included using convenience sampling. This sample size was determined based on previous ICU workload studies and statistical power analysis to ensure adequate data for evaluating NAS and patient outcomes. Participants were recruited over a one-year period, from 07 March 2022 to 27 February 2023. The Nursing Activities Score (NAS) was used to assess nursing workload per patient. NAS includes 23 nursing interventions; each assigned a specific weight based on the time required for completion. The total NAS score (ranging from 0 to 177%) reflects the percentage of a nurse’s time allocated to a single patient over 24 hours. Higher NAS values indicate increased nursing workload and resource demands. Outcome of patients were categorized as survivors and non-survivors based on ICU discharge status. Data on patient demographics, clinical condition, length of ICU stay, and severity of illness were collected. All statistical analyses were conducted using SPSS version 25.0. Continuous variables (NAS scores, age, ICU stay duration) were presented as median and interquartile range (IQR) as data was not normal. Categorical variables (survival status) were expressed as frequencies and percentages. Receiver Operating Characteristic (ROC) Analysis was used to evaluate the predictive ability of NAS for patient outcomes. The area under the curve (AUROC), cut-off score, sensitivity, and specificity were calculated. Hosmer-Lemeshow Goodness-of-Fit Test was used to assess the accuracy of the NAS model in predicting patient mortality. A p-value >0.05 indicated a good model fit. Logistic Regression Analysis was performed to examine the relationship between NAS scores and mortality risk, with odds ratio (Exp(B)) and confidence intervals (CI) reported. Mann-Whitney U Test was used to compare NAS scores between survivors and non-survivors to determine statistical significance (p < 0.05 considered significant). The study was carried out after the approval of research ethics committee of university and Nepal health research council (Ref: LUC/REC/INT/111/2022 and NHRC/071/2022) and permission for data collection was granted by the study site (Ref: CMC-IRC/078/079-150). The written consent was obtained from each respondent or legally authorized representative (LAR). Strict confidentiality of information was maintained.

## 4. RESULTS

A total of 501 critically ill patients admitted in adult ICUs were enrolled. The age of the patients ranged between 18 to 86 years. The median age of the critically ill patient was 48.96 (IQR:65-30). Males comprised 306 (61.1%) and female comprised 195 (38.9%) of the total number of patients. Of the 501 patients, 185 (36.9%) died and 316 (63.1%) survived. The median NAS score recorded in the study was 83.40, with an interquartile range (IQR) of 101.10 – 68.30 and a range from 39.2 to 134.4. This result indicates a high nursing workload in the ICU, suggesting that nurses are required to provide substantial care per patient. A higher NAS score generally reflects increased patient dependency and resource utilization, reinforcing the need for adequate staffing levels to ensure optimal patient care. The wide range of scores suggests variability in patient acuity levels, with some requiring minimal interventions while others demand intensive nursing care. For Length of Stay (LOS), the median was 6 days, with an IQR of 5 to 8 days, meaning that the middle 50% of patients had a hospital stay within this range. The shortest recorded LOS was 3 days, whereas the longest was 18 days, highlighting differences in patient recovery or care needs. NAS model discrimination was assessed with area under ROC. The Area Under the Receiver Operating Characteristic (AUROC) curve was 0.838 (p < 0.001), indicating a high predictive accuracy of NAS for patient outcomes. A cut-off score of 90.40 was determined, with 73.5% sensitivity and 73.1% specificity, meaning that NAS is a strong tool for distinguishing between patients at high and low risk of mortality. These findings are comparable to other ICU-based studies where higher NAS scores were associated with worse patient outcomes, highlighting the potential of NAS in early identification of critically ill patients requiring intensive care interventions (Table 1 and Figure 1). NAS model calibration was assessed with the Hosmer-Lemeshow Goodness-of-Fit test. The Hosmer-Lemeshow test yielded a Chi-square value of 8.967 and a p-value of 0.422, indicating a good fit of the NAS model in predicting ICU patient outcomes. Since the p-value is greater than 0.05, there is no significant difference between observed and predicted mortality rates, confirming that NAS is a statistically reliable model for assessing nursing workload and patient risk (Table 2). This result validates NAS as a useful decision-making tool for ICU workload management, allowing nurse staffing levels and care intensity to be adjusted based on predicted patient risk levels. Predictive performance of NAS was assessed with Logistic Regression Analysis. The logistic regression analysis produced a B coefficient of -0.065 and an odds ratio (Exp(B)) of 0.937 (p < 0.001), indicating that higher NAS scores are significantly associated with increased mortality risk. The negative regression coefficient suggests that as NAS increases, the likelihood of survival decreases, supporting the idea that critically ill patients requiring more nursing interventions have a higher risk of adverse outcomes. The 95% confidence interval (0.926 – 0.948), which does not include 1, confirms that this relationship is statistically significant, further reinforcing the importance of workload measurement in ICU settings (Table 3). The comparison of median NAS scores between survivors and non-survivors revealed a significant difference (p < 0.001). Non-survivors had a median NAS of 110.70, while survivors had a median NAS of 76.20. The Mann-Whitney U test value (9464.00) further confirmed a statistically significant disparity in nursing workload between these two groups (Table 4). This result suggests that non-survivors required considerably more nursing interventions, reflecting their higher disease severity and poor outcomes, emphasizing the need for strategic workload distribution and improved ICU nurse staffing to manage critically ill patients effectively. Overall, the data analysis confirms that NAS is a reliable tool for assessing nursing workload and predicting patient outcomes. The NAS values indicate that ICU nurses have a significant workload, with considerable variation across patients. The findings demonstrate that higher NAS scores correlate with increased patient acuity and mortality, highlighting the need for adequate ICU staffing, workload distribution, and early intervention strategies. Given the statistical validity of NAS, healthcare facilities can integrate it into workforce planning and patient risk assessments, ensuring better patient outcomes and optimized resource allocation in ICU settings.

**Figure 1:**
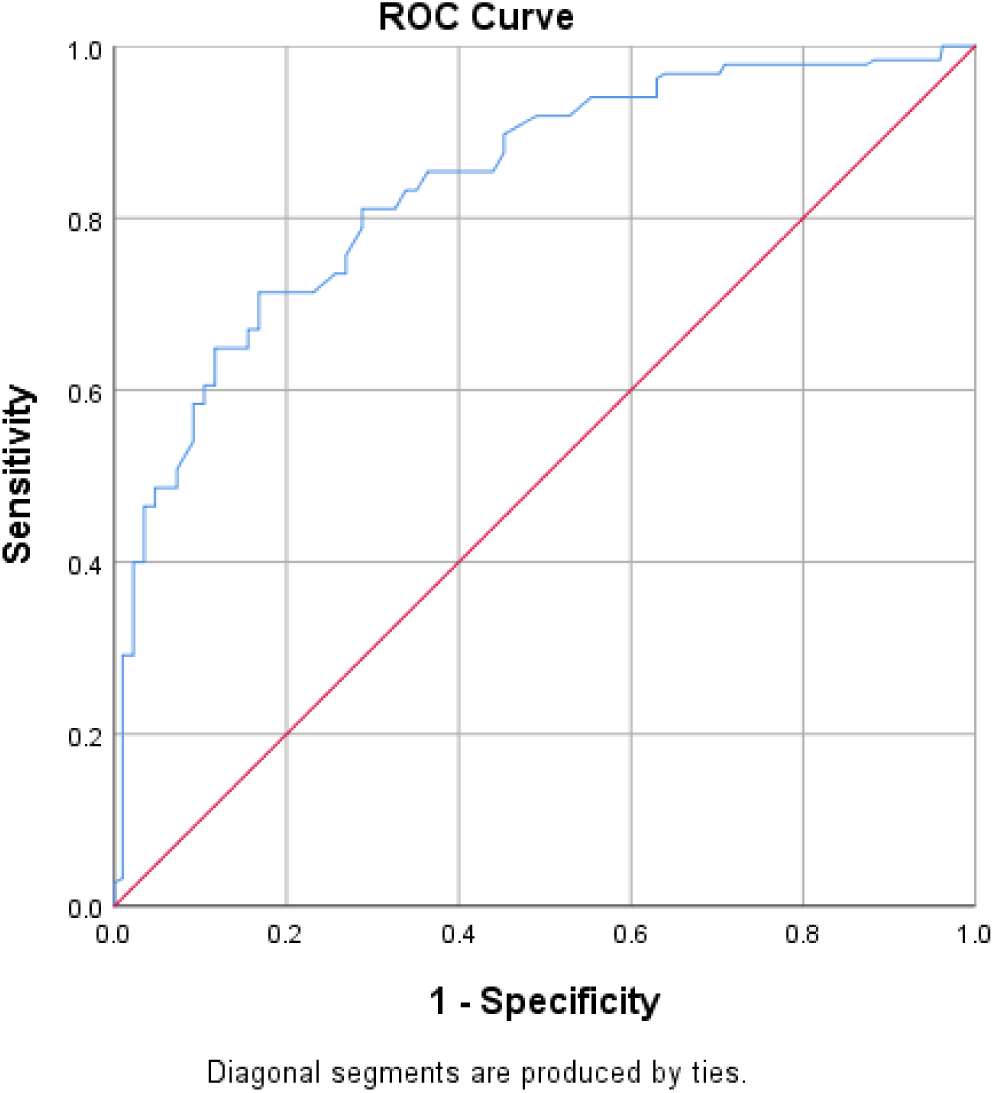
Area under Receiver Operating Characteristic curve (AUROC=0.838) for the Nursing activities score (NAS)

**Table 1:** Receiver Operating Characteristic curve (ROC) analysis for the Nursing activities score (NAS) Scoring System for outcome of critically ill patients.

| Scoring System | Cut off point | AUROC | p value | Sensitivity | Specificity | 95% CI |  |
| --- | --- | --- | --- | --- | --- | --- | --- |
|  |  |  |  |  |  | Lower | Upper |
| NAS | 90.40 | 0.838 | <0.001 | 0.735% | 0.731% | .802 | .875 |

**Table 2:** The Hosmer-Lemeshow test for the goodness-of-fit evaluation for Nursing activities score (NAS)

| Scoring System | H-L Chi-square | p-value |
| --- | --- | --- |
| NAS | 8.967 | 0.422 |

**Table 3:**
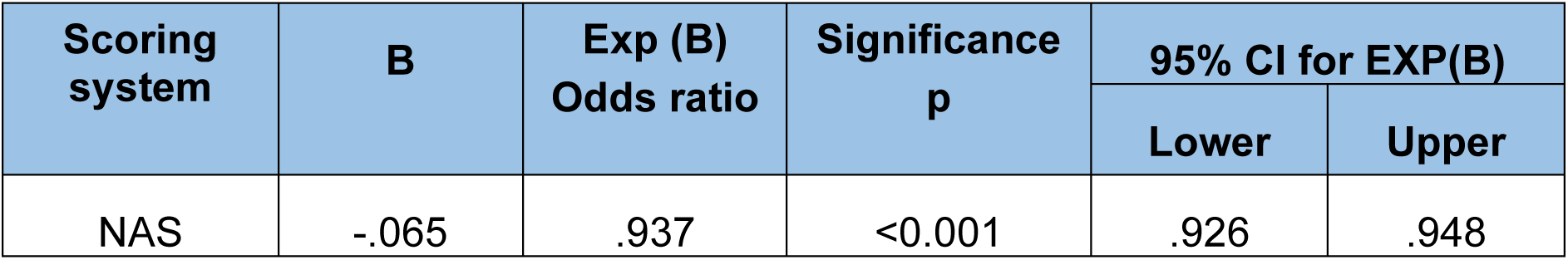
Predictive performance assessment for the NAS scoring system.

| Scoring system | B | Exp (B)<br>Odds ratio | Significance<br>p | 95% CI for EXP(B) |  |
| --- | --- | --- | --- | --- | --- |
|  |  |  |  | Lower | Upper |
| NAS | -.065 | .937 | <0.001 | .926 | .948 |

**Table 4:** Comparison of median nursing activities score (NAS) among Survivors and non-survivors.

| Scoring system | Number | Median | Mean Rank | Mann-Whitney U | p-value |
| --- | --- | --- | --- | --- | --- |
| **NAS** |  |  |  |  |  |
| Non-survivor | 185 | 110.70 | 357.84 | 9464.00 | <0.001 |
| Survivor | 316 | 76.20 | 188.45 |  |  |

## 5. DISCUSSION

This study assessed nursing workload in an ICU setting using the Nursing Activities Score (NAS) and evaluated its predictive value for patient outcomes. The findings revealed that higher NAS scores were associated with increased mortality risk, suggesting a strong correlation between workload, patient severity, and survival rates. These results align with previous studies highlighting the impact of nursing workload on patient care quality, ICU efficiency, and nurse well-being [1,10,13].

### Nursing Workload Assessment Using NAS

The median NAS score in this study was 83.40 (IQR: 101.10–68.30, range: 39.2–134.4), indicating high nursing workload in the ICU. This aligns with findings by Miranda et al. (2003), who reported that NAS scores exceeding 80% indicate an excessive workload, often requiring additional staffing and resource allocation [11]. Similar studies in ICUs across high-income countries have reported comparable NAS scores, reinforcing the global concern regarding ICU nursing workload [12,9,14].

High NAS scores indicate substantial nursing care requirements, particularly in critically ill patients who need continuous monitoring, invasive procedures, and mechanical ventilation. Alhosani MI et al. (2023) found that ICU patients requiring mechanical ventilation have significantly higher NAS scores, which aligns with our study’s findings [15]. The higher NAS scores in non-survivors suggest that critically ill patients require more intensive nursing care, a pattern consistent with studies linking workload intensity with disease severity [10,14].

### Predictive Value of NAS for Patient Outcomes

The Receiver Operating Characteristic (ROC) analysis demonstrated that NAS has strong predictive power for ICU patient outcomes, with an AUROC of 0.838 (p < 0.001). This suggests that NAS is an effective tool for identifying high-risk patients. The cut-off NAS score of 90.40, with 73.5% sensitivity and 73.1% specificity, indicates that patients requiring a NAS above this threshold are more likely to experience adverse outcomes. Similar findings have been reported in studies where higher NAS scores correlated with ICU mortality [14,16].

In a study by Nassiff A et al. (2018), NAS was found to be more accurate than traditional severity scoring systems, such as APACHE II and SOFA, in predicting mortality, reinforcing its value in ICU settings [17]. These findings support the argument that NAS should be integrated into ICU patient monitoring systems to improve early risk identification and workload distribution among nurses.

### Model Fit and Reliability of NAS

The Hosmer-Lemeshow goodness-of-fit test (Chi-square = 8.967, p = 0.422) confirmed that the NAS model fits well with observed ICU patient outcomes. A well-calibrated model strengthens the case for integrating NAS into ICU management strategies, as demonstrated in previous studies advocating for data-driven nursing workforce planning [3,18].

Similar findings were reported by Sardo PMG et al. (2023), where NAS demonstrated strong predictive accuracy across different ICU populations [16]. The strong statistical reliability of NAS suggests that it could be used as a workload monitoring tool, enabling hospitals to improve staff allocation, enhance patient safety, and reduce ICU strain.

### Association Between NAS and Mortality

The logistic regression analysis revealed a negative B coefficient (-0.065) and an odds ratio (Exp(B) = 0.937, p < 0.001), suggesting that higher NAS scores are linked to a lower probability of survival. This contradicts the assumption that higher nursing workload leads to better outcomes, instead emphasizing that increased NAS scores reflect greater patient severity rather than improved survival chances [9,4,19].

The comparison of NAS between survivors and non-survivors further supports this finding. Non-survivors had a median NAS of 110.70, significantly higher than survivors (76.20, p < 0.001). This is consistent with studies demonstrating that higher nursing workload is associated with critically ill patients who have a higher risk of mortality [10,12]. The Mann-Whitney U test results (p < 0.001) confirm a significant difference in workload between survivors and non-survivors, reinforcing the notion that ICU patients requiring intensive nursing care are at greater risk of mortality.

### Implications for ICU Staffing and Nursing Workload Management

The findings of this study highlight the urgent need for workforce optimization in ICUs, particularly in low-middle-income countries (LMICs) like Nepal. The high NAS scores indicate a substantial nursing workload, emphasizing the necessity of maintaining an adequate nurse-to-patient ratio [7]. Studies have shown that higher nurse staffing levels lead to better patient outcomes and reduced ICU mortality rates [1,4].

Additionally, Ghabi AAH et al. (2024) found that increasing the number of registered nurses per shift significantly reduced adverse events, highlighting the need for evidence-based staffing policies [13]. Future studies should evaluate real-time workload balancing using NAS scores, as proposed by Hasselgård AM et al. (2024), to improve ICU efficiency and patient safety [14].

This study confirms that NAS is a reliable tool for measuring nursing workload and predicting ICU patient outcomes. Higher NAS scores were associated with increased mortality risk, reflecting greater patient severity and resource needs. The findings highlight the critical role of adequate staffing in ICUs, emphasizing the need for data-driven workforce planning to balance workload distribution and optimize patient care.

By integrating NAS-based workload assessments into ICU staffing strategies, hospitals can enhance nurse efficiency, reduce burnout, and improve patient survival rates. Future research should explore multi-center validation of NAS and its application in diverse ICU settings, particularly in low-resource environments, to develop evidence-based staffing models for critical care.

### Strengths, Limitations, and Nursing Implications

This study has notable strengths. It is the first to systematically assess nursing workload in intensive care units in Nepal using the Nursing Activities Score (NAS), thereby addressing a significant gap in local and regional evidence. The relatively large sample of critically ill patients and the application of robust statistical analyses, including ROC curve, logistic regression, and model fit testing, provide strong validity to the findings. By comparing survivors and non-survivors, the study demonstrates the direct clinical relevance of nursing workload to patient outcomes in resource-limited ICU settings.

However, several limitations should be acknowledged. Conducting the study in a single teaching hospital limits generalizability to other hospitals and regions. As an observational design, the study cannot establish causality between workload and patient outcomes. Reliance on NAS alone without incorporating other workload measures (e.g., nurse burnout, skill mix, or organizational factors) may not fully capture the complexity of ICU care. In addition, data were collected during particular period of time and may not reflect workload variations across shifts or seasonal patterns. Unmeasured confounders, such as physician practices and availability of resources, may also have influenced patient outcomes.

These findings have important clinical and policy implications. Future research should expand to multi-center and longitudinal designs, incorporating broader measures of workload and contextual factors. From a practical standpoint, NAS could be adopted as a routine monitoring and workforce planning tool to identify high-risk patients and allocate nursing resources more effectively. Improving nurse-to-patient ratios, guided by NAS assessments, may reduce preventable adverse events, improve patient safety, and promote sustainable working conditions for nurses in intensive care settings of low- and middle-income countries.

## CONCLUSION

The study assessed nursing workload and the utility of the Nursing Activities Score (NAS) in predicting outcomes for critically ill ICU patients. The results demonstrate that patients who did not survive required substantially more nursing care, as reflected by higher NAS scores. The NAS tool effectively distinguished between survivors and non-survivors, indicating its strength as a predictor of patient outcomes. Model evaluation confirmed that NAS provides a reliable fit for outcome prediction, and further analysis affirmed that increases in nursing workload were closely associated with a greater likelihood of mortality. These findings underscore the importance of using NAS not only to monitor nursing workload but also to identify patients at higher risk of poor outcomes. Overall, the study supports the clinical relevance of NAS as a practical and informative tool for guiding nurse staffing decisions and enhancing patient care in intensive care settings.

## AUTHORS’ CONTRIBUTIONS

RKM: Conceptualization, design of the work, data collection, data curation, formal analysis, manuscript draft, review and editing manuscript

HCH: Conceptualization, formal analysis, supervision, review, editing of manuscript

BB: Conceptualization, formal analysis, supervision, review, editing of manuscript

MSN: data collection, formal analysis, writing, review, editing of manuscript

All authors read and approved the final manuscript.

## LIST OF ABBREVIATIONS

AUROC: Area Under Receiver Operating Characteristics Curve
IQR: Interquartile range
LUC: Lincoln University College
REC: Research Ethics Committee
INT: International
CMC-IRC: Chitwan Medical College-Institutional Review Committee
Sens: Sensitivity
Spec: Specificity
L: Lower
U: Upper
CI: Confidence Interval
H-L: Hosmer-Lemeshow test

## ETHICS APPROVAL AND CONSENT TO PARTICIPATE

The study was carried out after the approval of research ethics committee of university and Nepal health research council (Ref: LUC/REC/INT/111/2022 and NHRC/071/2022) and permission for data collection was granted by the study site (Ref: CMC-IRC/078/079-150). The written consent was obtained from each respondent or legally authorized representative (LAR). Strict confidentiality of information was maintained.

## AVAILABILITY OF DATA AND MATERIALS

The data supporting the findings of this article are available from the corresponding author upon reasonable request.

## FUNDING

This study was conducted with the authors’ personal funding.

## CONFLICT OF INTEREST

The authors declare that this study forms part of the corresponding author’s PhD research at Lincoln University College, Malaysia, and was conducted at Chitwan Medical College Teaching Hospital, Nepal. The work was carried out in accordance with University academic requirements for doctoral research. The authors report no financial, personal, or other conflicts of interest related to this study.

## ACKNOWLEDGEMENTS

The authors would like to acknowledge the support of all study participants, ICU In charge, ICU staffs, Prof. Dr. Harishchandra Neupane (Chairman and Managing Director-CMC), Prof. Dr. Gopendra Prasad Deo (HOD, Department of Anesthesiology and Critical Care Medicine-CMC) and Assoc. Prof. Jaya Prasad Singh for statistical analysis.

## Notes

### Clinical Trial

"N/A"

### Funding Statement

This study was conducted with the authors' personal funding. No funding received from any funding organization.

### Author Declarations

The study was carried out after the approval of the research ethics committee of the university and the Nepal Health Research Council (Ref: LUC/REC/INT/111/2022 and NHRC/071/2022) and permission for data collection was granted by the study site (Ref: CMC-IRC/078/079-150). The written consent was obtained from each respondent or legally authorized representative (LAR). Strict confidentiality of information was maintained. Data confidentiality was maintained, and patient identities were anonymized. All human research procedures followed were in accordance with the ethical standards of the committee responsible for human experimentation (institutional and national) and with the Helsinki Declaration of 1975, as revised in 2013. Participation was voluntary, and individuals had the freedom to withdraw at any point without facing any consequences.

